# Early life PM_2.5_ exposure, childhood cognitive ability and mortality between age 11 and 86: A record-linkage life-course study from Scotland

**DOI:** 10.1101/2023.05.18.23289845

**Authors:** Gergő Baranyi, Lee Williamson, Zhiqiang Feng, Sam Tomlinson, Massimo Vieno, Chris Dibben

## Abstract

**Background:** Living in areas with high air pollution concentrations is associated with all-cause and cause-specific mortality. Exposure in sensitive developmental periods might be long-lasting but studies with very long follow-up are rare, and mediating pathways between early life exposure and life-course mortality are not fully understood.

**Methods:** Data were drawn from the Scottish Longitudinal Study Birth Cohort of 1936, a representative record-linkage study comprising 5% of the Scottish population born in 1936. Participants had valid age 11 cognitive ability test scores along with linked mortality data until age 86. Fine particle (PM_2.5_) concentrations estimated with the EMEP4UK atmospheric chemistry transport model were linked to participants’ residential address from the National Identity Register in 1939 (age 3). Confounder-adjusted Cox regression estimated associations between PM_2.5_ and mortality; regression-based causal mediation analysis explored mediation through childhood cognitive ability.

**Results:** The final sample consisted of 2734 individuals with 1608 deaths registered during the 1,833,517 person-months at risk follow-up time. Higher early life PM_2.5_ exposure increased the risk of all-cause mortality (HR=1.03, 95% CI: 1.01-1.04 per 10μg m^-3^ increment), associations were stronger for mortality between age 65 and 86. PM_2.5_ increased the risk of cancer-related mortality (HR=1.05, 95% CI: 1.02-1.08), especially for lung cancer among females (HR=1.11, 95% CI: 1.02-1.21), but not for cardiovascular and respiratory diseases. Higher PM_2.5_ in early life (≥50μg m^-3^) was associated with lower childhood cognitive ability, which, in turn, increased the risk of all-cause mortality and mediated 25% of the total associations.

**Conclusions:** In our life-course study with 75-year of continuous mortality records, we found that exposure to air pollution in early life was associated with higher mortality in late adulthood, and that childhood cognitive ability partly mediated this relationship. Findings suggest that past air pollution concentrations will likely impact health and longevity for decades to come.

**HIGHLIGHTS:**

- We explored PM_2.5_ at age 3 and mortality between age 11 and 86 in a Scottish cohort
- PM_2.5_ increased the risk of all-cause mortality, especially between the age of 65 and 86
- Childhood cognitive ability mediated 25% of the total association
- Associations were prominent for (lung) cancer mortality, especially among females
- Air pollution in early life may affect health and longevity across the life course

## INTRODUCTION

Ambient air pollution poses one of the greatest environmental threats to human health. Fine particulate matter with the aerodynamic diameter of less than 2.5µm (PM_2.5_) is the fifth largest mortality risk factor, with an estimated 4.2 million attributable death and 103 million disability adjusted life years in 2015 (1). Consistent and of high certainty evidence links PM_2.5_ exposure to increased risk of all-cause and cause-specific (e.g., cardiovascular, lung cancer, respiratory) mortality in the general population (2–5). Large number of studies focussed on the short (from one hour to 7 days) (5), medium (<10 years) and long-term impacts (<25 years) (2) or poor air quality, however, the number of investigations drops when it comes to very-long term (≥25 years) associations (6–11).

Exposure to high levels of air pollution may be particularly detrimental and long-lasting when it intersects with sensitive periods of human development; still, few studies followed up narrow-age cohorts (9) which is crucial to assess the developmental timing of exposure. Evidence suggests that the time before birth and the first years of life are critical, providing the foundation of later health and lifespan development (12, 13). Air pollution exposure around birth is associated with adverse birth outcomes (14, 15), altered brain development (16), and modifications in DNA methylation patterns (17), key predictors of healthy ageing and longevity (18, 19). It is plausible that air pollution during this early sensitive/critical period has very long-term impact on health (and subsequent mortality), likely lasting throughout the entire life course. Still, due to lack of historical air pollution data and cohort studies with life-course follow-up, previous investigations were not able to explore this hypothesis.

Using a representative cohort of the Scottish population born in 1936, we examined whether and how exposure to PM_2.5_ in early life contributed to a) all-cause mortality across almost the entire life course (i.e., between age 11 and 86), to b) cause-specific mortality during late adulthood, and c) whether associations with all-cause mortality were mediated by childhood general cognitive ability.

## METHODS

Data were taken from the Scottish Longitudinal Study Birth Cohort of 1936 (SLSBC1936), a retrospective cohort study assembled from routinely collected administrative data, representative of the Scottish population born in 1936. SLSBC1936 is based on very high-quality data linkages between four sources, a) the 1939 National Identity Register, a census-like registry of the entire UK population carried out on the 29^th^ September 1939 at the start of World War Two; b) the Scottish Mental Survey 1947 (SMS1947), a general cognitive ability test undertaken among almost all 11-year old Scottish schoolchildren in 4^th^ June 1947; c) Scotland’s National Health Services Central Register (NHSCR); and d) the Scottish Longitudinal Study, a 5.3% administrative sample of the Scottish population selected based on 20 semirandom birthdays and followed up since 1991 (20). In addition to the core SLSBC1936 sample, we also included individuals who would have entered the Scottish Longitudinal Study if they did not die or leave the country before in 1991; detailed information on matching procedure are available elsewhere (20). Therefore, our sample comprised individuals born in 1936 on one of the 20 Scottish Longitudinal Study birth dates, enumerated in the 1939 National Identity Register, participated in the SMS1947, and captured by the NHSCR system, totalling to a maximum sample size of 3626 individuals.

### Exposure to PM_2.5_ pollution

Annual level of PM_2.5_ concentrations for the modelling year of 1935 was estimated using the EMEP4UK version 4.17 derived from the EMEP MSC-W version 4.17 (21) atmospheric chemistry transport model (22–24). The meteorology year used here was 2015. The meteorological driver used was the Weather Research and Forecast (WRF) model version 3.9.1.1 (25) based on the Global forecast system final re-analysis (GFS-FNL) as initial condition and 6-h nudging (26). Gridded anthropogenic emission of pollutants (nitrogen oxides, sulphur oxides, ammonia, non-methane volatile organic compounds, carbon monoxide, course and fine particulate matter) were estimated for the year 1950 (27), and a scaled version of these data distributions, based upon activity data research, were used to make emission estimates in 1935 (28). Non-NH_3_ activity data in that era are largely a reflection of the use of fossil fuels such as coal and of some oil-derived products while agricultural data, the majority source of NH_3_ emissions, was derived from the Vision of Britain database (29). The model employs a horizontal resolution of 0.5 x 0.5 degrees to a greater European domain providing the boundary condition for a nested UK domain with a horizontal resolution of 0.037 x 0.037 degrees (∼ 3km × 4km). Estimated PM_2.5_ concentrations ranged between 4.2μg m^-3^ to 116.9μg m^-3^ for Scotland (Figure 1). The EMEP4UK model has been evaluated in the UK and internationally showing good agreement between modelled and observed concentrations (30, 31); historical concentrations of air pollution exposure derived from this model have been used in previous epidemiological studies (12, 28).

**Figure 1:**
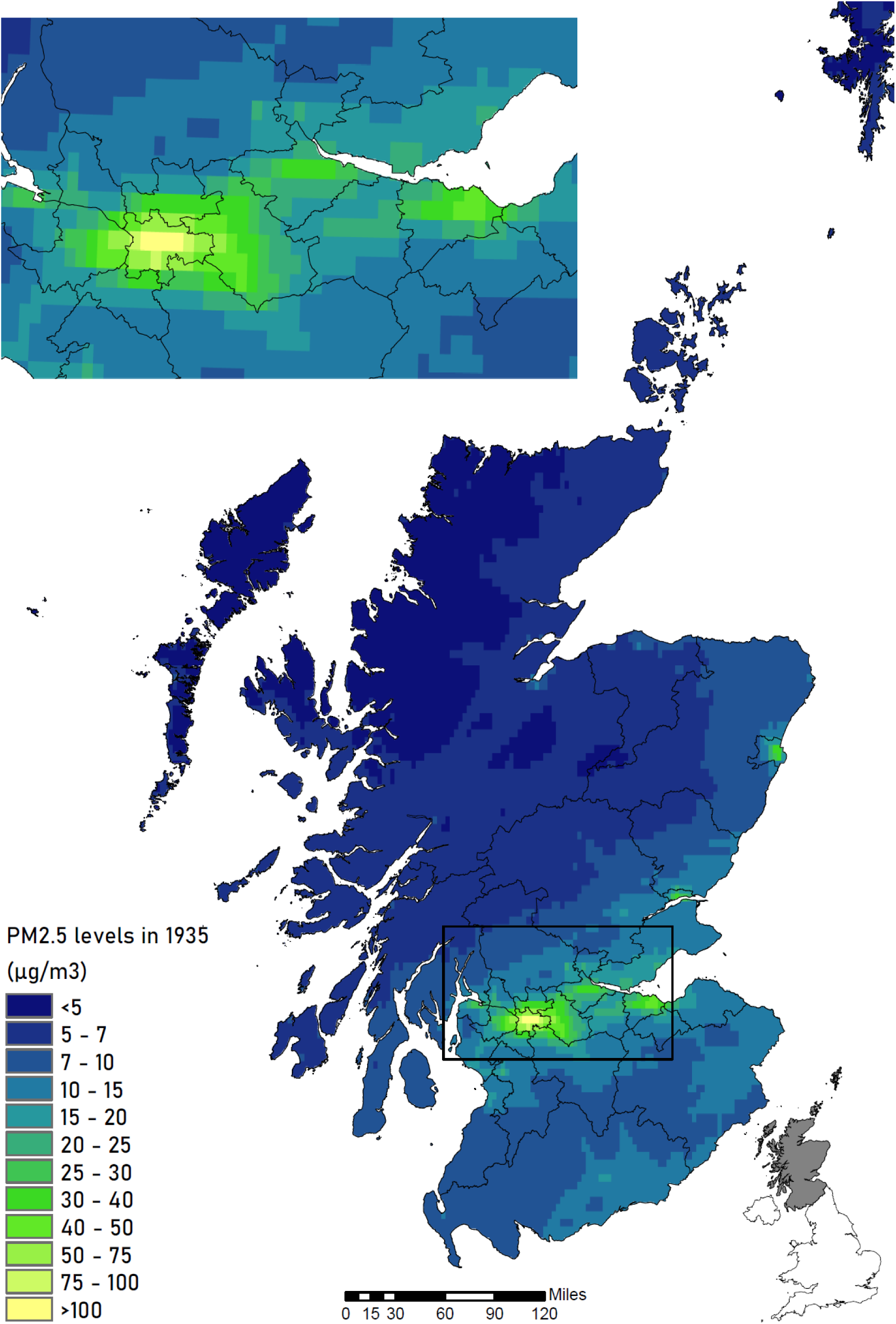
Mean 1935 PM_2.5_ concentration (in μg m^-3^) in Scotland, and in the Central Belt of Scotland, estimated using the EMEP4UK atmospheric chemistry transport model with a horizontal resolution of ∼3 × 4 km. Index map in the right bottom corner presents Scotland’s location within the United Kingdom; black lines are council areas based on current boundaries.

Participants’ residential addresses were transcribed from the 1939 National Register and were geocoded using the Historical Address Geocoding – GIS software (32), which links historical records to contemporary addresses. Grid reference was linked to the sample using the 1939 street index (records with house numbers, street names, enumeration district and the registration district codes) and centroid of the 1939 enumeration districts, with very high linkage rate (33). Geocoded 1939 addresses were intersected with PM_2.5_ concentrations modelled for the year 1935.

### Mortality records

All-cause mortality was derived from NHSCR data indicating the month and year of death between June 1947 (i.e., date of SMS1946) and June 2022 (i.e., 901 months). The NHSCR data also includes information for those deaths occurring in other parts of the United Kingdom or overseas, which are not part of the routine Vital Event deaths (i.e., civil registration deaths). Cause-specific mortality was obtained from the routine Vital Events deaths database, available only for those who became part of the Scottish Longitudinal Study after 1991 and died within Scotland between April 1991 (i.e., date of 1991 Census) and December 2017 (i.e., 321 months). Cause of death was reported according to the International Classification of Diseases 9^th^ Revision (ICD-9) or 10^th^ Revision (ICD-10) which we classified into natural causes (ICD-9: 001 to 799; ICD-10: A00 to R99), cardiovascular (ICD-9: 390 to 459; ICD-10: I00 to I99), respiratory (ICD-9: 460 to 519; ICD-10: J00 to J99), and cancer-related deaths (ICD-9 140 to 239; ICD-10: C00 to D48).

### General cognitive ability in childhood

On the Wednesday 4^th^ June 1947, 94% of all Scottish children born in 1936 (n=70805) participated on a cognitive assessment as part of the SMS1947. The survey utilised the Moray House No.12 test, a 71 item general cognitive ability test, including mental tasks such as same-opposites, word classification, analogies, reasoning, cipher decoding and spatial items (18, 34). Tests were administered in school classes across the entire country on the same day, with the same instructions and within a 45-minute test interval (18). The test score had a maximum value of 76 and it correlated highly with the Stanford-Binet test providing good criterion validity (34).

### Covariates

Potential confounders of the exposure, outcome and mediator relationship were identified from the literature and are presented in a directed acyclic graph (Figure 2). Sex (male, female) and age (in years) were derived from the SMS1946. Mother’s age (in years), mother’s marital status (married, not married) and parental occupational social position was derived from the 1939 National Identity Register. For the latter, occupational titles were first manually coded for each member of the household using the Historical International Classification of Occupations coding system (35). Occupational codes were assigned a value of occupational social position based on the HISCAM scale (36), which is a historic measure of occupational social stratification during the 19th and early 20th centuries. The continuous HISCAM score is centred around the mean of 50 with standard deviation of 10 in the underlying population; values may range between 1 and 99 with greater numbers indicating higher social position (36). To each participant, we assigned the father’s social position (or the mother’s if fathers’ not available); otherwise, we calculated the average social position of all other household members.

**Figure 2:**
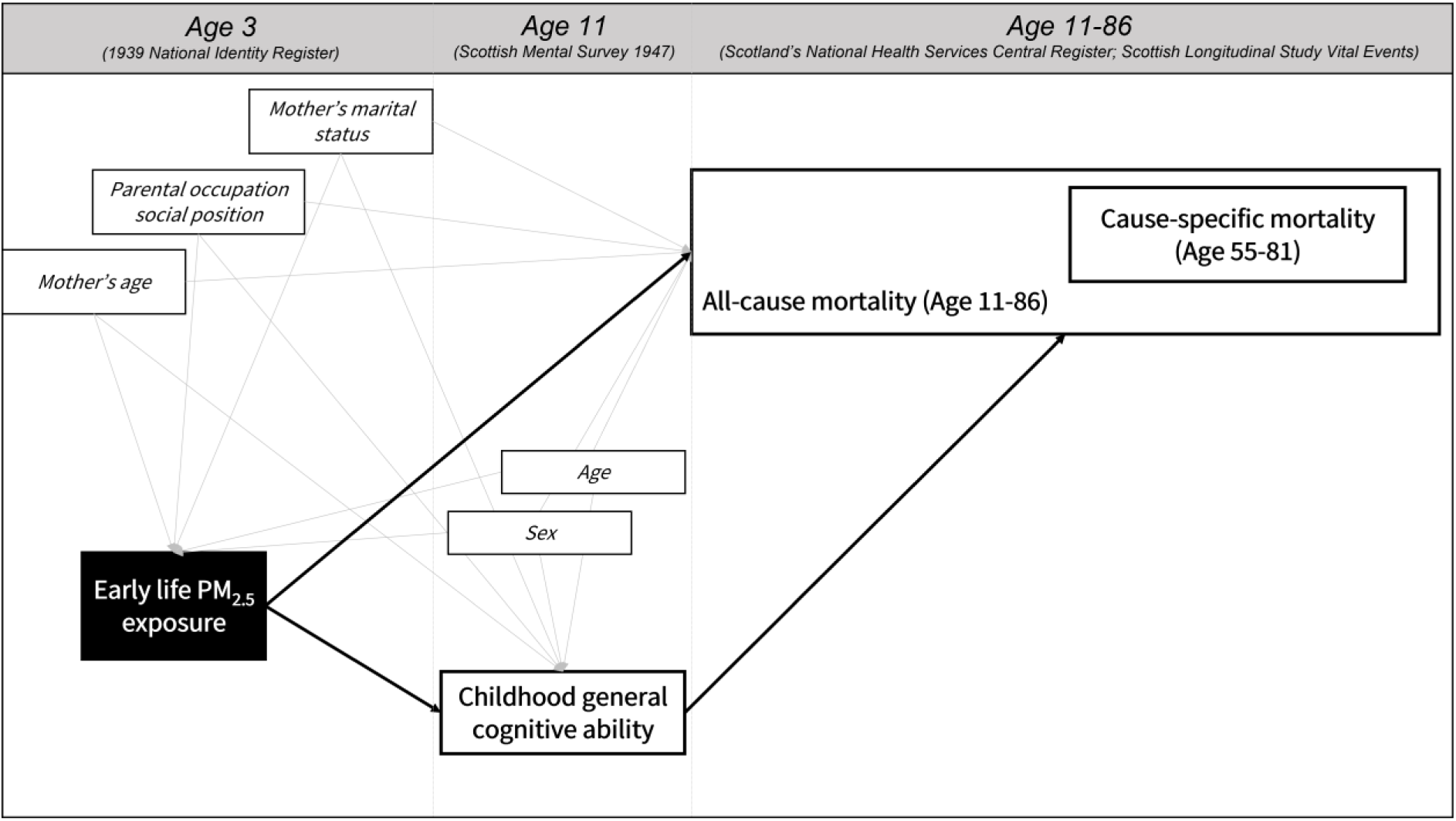
Directed acyclic graph presenting associations between exposure, mediator, outcome and confounders, and the main source of data. Associations between confounders are not shown for simplicity, solid black lines are associations of interest. Geocoded 1939 addresses were linked to PM_2.5_ concentrations estimated with the EMEP4UK model for the year 1935.

### Statistical analysis

Cox proportional hazards regressions estimated the associations between PM_2.5_ and mortality using months of follow-up as the time scale. Effect estimates were presented as Hazard Ratios (HR) with 95% confidence intervals (CI) for each 10μg m^-3^ incremental increase. Subjects were censored at the time of their death, at the time they last left Scotland (from NHSCR data), or at the end of the study period if no events were registered during the follow-up period. Based on the Schoenfeld residuals, the proportional hazards assumption was met for all covariates but for sex; therefore, we incorporated strata for sex and presented stratified findings for males and females. To determine the representativeness of the analytical sample, we compared the sex distribution, average age and average Moray House test scores with the population values coming from the SMS1947 (37).

The main analysis focussed on early life PM_2.5_ exposure and all-cause mortality. Two confounder models were set *a priori*: Model 1 controlled for age and sex (strata); Model 2 further adjusted for parental occupational position, mother’s age, and mother’s marital status. Continuous variables were scaled to support interpretation. In addition to treating the associations as time-independent, we also tested whether the relationship between PM_2.5_ exposure and all-cause mortality changed during follow-up by splitting the follow-up time into four intervals: first 527 months (i.e., average age of 11-55 years), from 528 to 647 months (i.e., average age of 55-65 years), from 648 to 767 months (i.e., average age of 65-75 years), and from 768 to 901 months (i.e., average age of 75-86 years). Finally, based on Model 2 adjustments we explored concentration-response relationship by adding penalized splines with 2, 3 and 4 degrees of freedom. The best model was determined based on the Bayesian Information Criterion (BIC) and linearity of relationship was examined with *χ*^2^ test.

Associations between PM_2.5_ in early life and mortality due to natural causes, as well as cardiovascular, respiratory and cancer mortality were estimated in competing risks analysis. Models were operationalised in Cox proportional hazards regression by fitting two endpoints at the same time (i.e., outcome of interest versus other deaths) with distinct baseline hazards for PM_2.5_, age, and sex (strata), and shared coefficients for parental occupational social position, mother’s age, and mother’s marital status. In contrary to calculating cause-specific hazards regression, this approach takes into consideration that competing events are mutually exclusive (38). Model 1 and Model 2 adjustments, as well as sex-stratified findings were presented.

Total association between PM_2.5_ exposure and all-cause mortality was decomposed into direct and indirect (through childhood general cognitive ability) effects applying regression-based causal mediation analysis within the counterfactual framework (39). For the mediator model, we fitted a linear regression, for the outcome model a Cox regression; the same Model 2 confounders were included in both mediator and outcome models. Percentage mediated was expressed as the percentage of total association explained by the mediator; 95% CIs for total, direct and indirect effects were estimated based on 10000 bootstrap replications. For the mediation analysis, PM_2.5_ exposure was dichotomised as <50 versus ≥50μg m^-3^, a cut off separating the highest ∼20% from the rest of the sample. We presented findings for the non-stratified sample only.

Six sets of sensitivity analyses were carried out. First, we presented main results (i.e., all-cause mortality) for alternative exposure operationalisations (i.e., interquartile range [IQR] increase, threshold [<50, ≥50μg m^-3^]). Second, we provided main models after adjusting for urban versus rural area of residence, which was determined based on living in Aberdeen, Dundee, Edinburgh, or Glasgow (i.e., urban) versus in other areas (i.e., rural), by intersecting 1939 addresses with current council boundaries. Third, proportion of missing data was significant for some of the covariates, therefore, we reran main models with multiple imputation by chained equation. Number of imputations was determined automatically based on the proportion of incomplete cases (40). Four, we dropped individuals with extremely high PM_2.5_ values (≥100μg m^-3^) residing in the centre of Glasgow City and reran main models. Five, instead of fitting two endpoints in Cox regression, competing risks were calculated with the Fine-Gray proportional subdistribution hazards regression. Last, we presented the mediation analysis with an alternative cut-off for ‘low’ and ‘high’ air pollution levels (<40 versus ≥40μg m^-3^) capturing ∼75% versus ∼25% of the sample.

Analyses were carried out using the *survival* (41), *regmedint* (42), *mice* (43) and *tidycmprsk* (44) packages in R 4.2.2 (45).

## RESULTS

The final SLSBC1936 sample without missing data included 2734 individuals (48.4% females) contributing to a total of 1,833,517 person-months (152,793.1 person-years) at risk with a median follow-up time of 771 months (64.3 years). During follow-up, 1608 individuals died, 475 left the study and 651 were still alive at the end of the study. The average exposure to PM_2.5_ was 31.3μg m^-3^ (SD=32.6; IQR=9.3-39.0). Comparing the SLSBC1936 analytical sample with the population of individuals born in 1936 (i.e., the main SMS1947 sample) suggested that neither the percentage of males and females, nor the average Moray House test scores differed between the SLSBC1936 sample and the population; however, our sample was approximately one month older than the population average likely due to the distribution of 20 semirandom birth days across the year (Table 1).

**Table 1:** Characteristics of participants in the analytical sample and in the respective population.

| Characteristics | Data source | Analytical sample<br>(SLSBC1936)<br>(n=2734) | Population<br>(SMS1947)<br>(n=70805)<sup>a</sup> | p-value<sup>b</sup> |
| --- | --- | --- | --- | --- |
| PM<sub>2.5</sub> exposure in early life in $\mu\text{g m}^{-3}$, mean ± SD | Age 3 | 31.3 ± 32.6 | NA | NA |
| Parental occupational social position, mean ± SD | (1939NIR, 28<sup>th</sup> | 53.8 ± 9.4 | | |
| Mother's marital status, n (%) | September |  |  |  |
| Married | 1939) | 2645 (96.7%) |  |  |
| Not married |  | 89 (3.3%) |  |  |
| Mother's age in years, mean ± SD | | 32.1 ± 6.4 | | |
| Age in years at the Moray House test, mean ± SD | Age 11 | 11.0 ± 0.3 | 10.9 ± 0.3<sup>c</sup> | <0.001 |
| Sex, n (%) | (SMS1947, 4<sup>th</sup> |  |  |  |
| Female | June 1947) | 1322 (48.4%) | 34996 (49.4%) |  |
| Male |  | 1412 (51.6%) | 35809 (50.6%) | 0.915 |
| Moray House test score, mean ± SD | | 36.8 ± 15.5 | 36.7 ± 16.1 | 0.940 |

Source: Scottish Longitudinal Study. Abbreviation: NA=not applicable; SD=standard deviation; SLSBC1936=Scottish Longitudinal Study Birth Cohort of 1936; SMS1947=Scottish Mental Survey 1947; 1939NIR=1939 National Identity Register.
<sup>a</sup> Reference values are from (37).
<sup>b</sup> P-values are based on one-sample t-test for mean difference and $\chi^2$ tests for difference in distribution.
<sup>c</sup> Own calculation based on Table IV (page 82 (37))

After adjusting for age and sex (strata), we found 3% higher hazard of all-cause mortality per 10μg m^-3^ increment increase in early life PM_2.5_ exposure (HR=1.03, 95% CI: 1.01-1.04). The association remained the same after further adjustments for parental occupation social position, mother’s marital status, and mother’s age (HR=1.03, 95% CI: 1.01-1.04) (Table 2). Associations were varying across the life course: while between age 11 and 65 there was no association, 10μg m^-3^ higher early life PM_2.5_ exposure increased the hazard of all-cause mortality between age 65 and 75 years by 4% (95% CI: 1.02-1.07), and between age 75 and 86 by 3% (95% CI: 1.00-1.05) (Model 2). Stratified analyses suggested that among females the association was strongest between age 75-86, while among males it was strongest between age 65-75 (Table 2). Furthermore, in the total sample, concentration-response relationship was monotonic, linear and best-fitting with two degrees of freedom (likelihood ratio test for non-linear component: *χ^2^*=0.43; *p*=0.530; see Bayesian Information Criterion values in Supplementary Table 1). HRs constantly increased with increasing PM_2.5_ concentrations but first crossed the value of HR=1.0 at the value of ∼38μg m^-3^. Although the shape of the association visually differed between females and males, confidence intervals were wide and overlapping (Figure 3).

**Figure 3:**
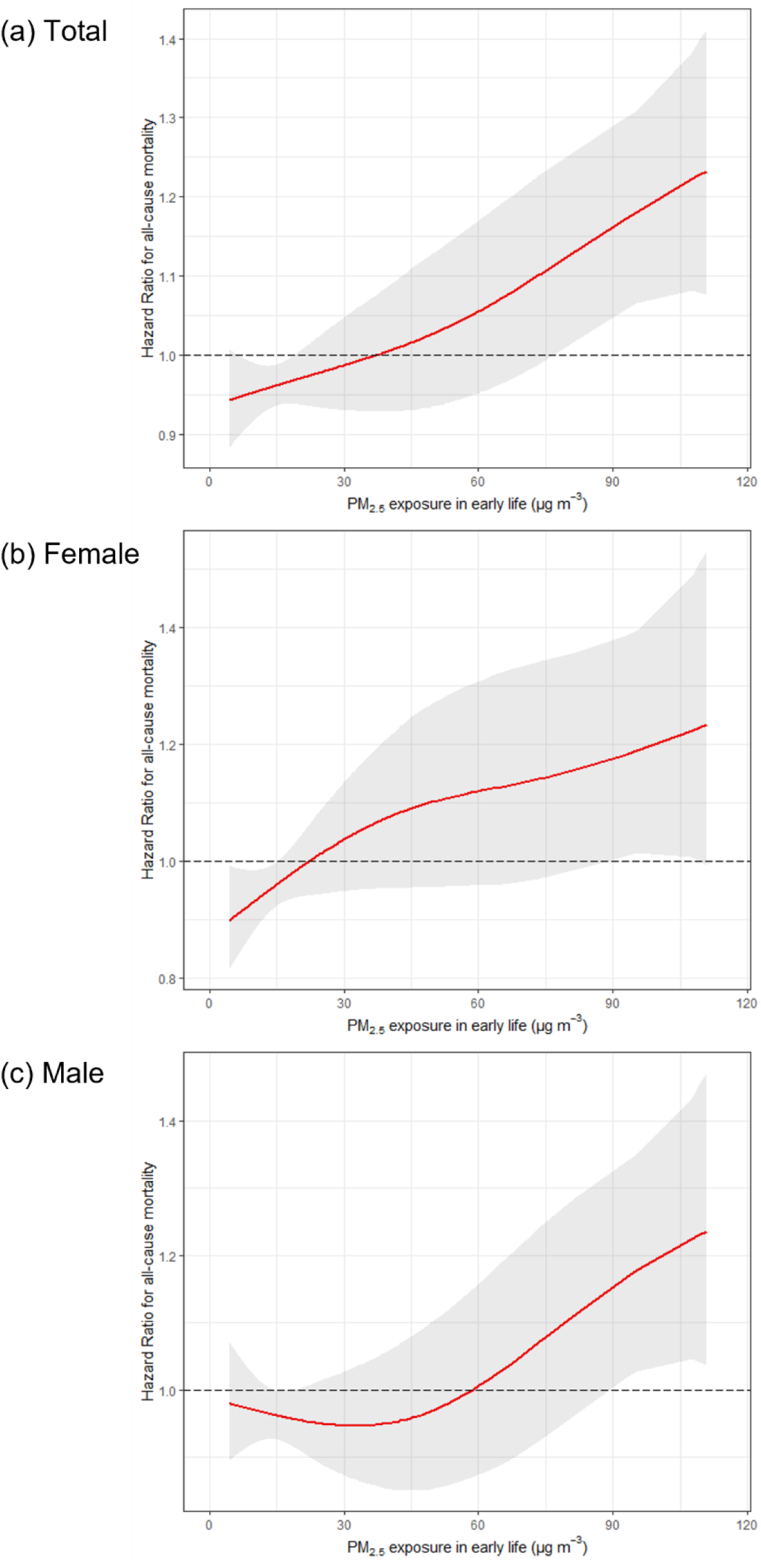
Concentration-response relationship between PM_2.5_ in early life and all-cause mortality in the Scottish Longitudinal Study Birth Cohort of 1936 for the (a) total sample; and among (b) females and (c) males. Curves were obtained by penalised splines with two degrees of freedom; observed relationships were linear. All-cause mortality was available between June 1947 and June 2022. Model was adjusted for age, (sex (strata),) parental occupational social position, mother’s marital status, and mother’s age. The total sample size was *n*=2734. Source: Scottish Longitudinal Study.

**Table 2:** Associations between early life PM_2.5_ exposure and all-cause mortality in the Scottish Longitudinal Study Birth Cohort of 1936 a) during the total follow up and b) by follow-up intervals.

| Covariates | Model 1<br>HR (95% CI) | Model 2 |  |  |
| --- | --- | --- | --- | --- |
|  |  | Total<br>HR (95% CI) | Female<br>HR (95% CI) | Male<br>HR (95% CI) |
| *a) During total follow-up* |  |  |  |  |
| PM<sub>2.5</sub> in early life (10µg m<sup>-3</sup> increase) | **1.03 (1.01-1.04)** | **1.03 (1.01-1.04)** | **1.03 (1.01-1.05)** | **1.02 (1.00-1.04)** |
| Age (per 1 SD increase) | 1.01 (0.96-1.06) | 1.01 (0.96-1.06) | 1.00 (0.93-1.07) | 1.02 (0.96-1.09) |
| Parental occupational social position (per 1 SD increase) |  | **0.92 (0.87-0.97)** | **0.91 (0.84-0.99)** | **0.92 (0.85-0.99)** |
| Mother's marital status |  |  |  |  |
| Married |  | ref | ref | ref |
| Not married |  | 1.22 (0.93-1.60) | 1.07 (0.73-1.57) | 1.42 (0.96-2.08) |
| Mother's age (per 1 SD increase) |  | 0.96 (0.91-1.01) | 0.96 (0.89-1.03) | 0.96 (0.90-1.03) |
| *b) By follow-up intervals<sup>a</sup>* |  |  |  |  |
| PM<sub>2.5</sub> in early life (10µg m<sup>-3</sup> increase) |  |  |  |  |
| Age 11-55 year | 1.01 (0.97-1.05) | 1.00 (0.96-1.04) | 1.01 (0.94-1.08) | 1.00 (0.95-1.05) |
| Age 55-65 year | 1.01 (0.97-1.05) | 1.01 (0.97-1.05) | 1.05 (0.99-1.12) | 0.99 (0.94-1.03) |
| Age 65-75 year | **1.05 (1.02-1.08)** | **1.04 (1.02-1.07)** | 1.02 (0.97-1.06) | **1.06 (1.03-1.10)** |
| Age 75-86 year | **1.03 (1.00-1.05)** | **1.03 (1.00-1.05)** | **1.04 (1.01-1.07)** | 1.02 (0.98-1.05) |

Source: Scottish Longitudinal Study. The sample size was $n=2734$ . Cox proportional hazard regression were fitted and Hazard Ratios (HR) with 95% confidence intervals (CI) are presented. In the total sample, sex as strata was included. Bolded values are significant ( $p<0.05$ ). All-cause mortality was available between June 1947 and June 2022. Abbreviation: SD=standard deviation.
<sup>a</sup> Covariates are not shown.

The association with cause-specific mortality was assessed in a restricted sample (*n*=1947) as information on cause of death was only available between April 1991 and December 2017. During this period 939 deaths were registered due to natural causes, 321 cardiovascular, 109 respiratory, and 341 cancer-related deaths. Competing risks analysis with adjustment for all covariates suggested that early life PM_2.5_ exposure increased the hazard of cancer-related mortality by 5% (95% CI: 1.02-1.08) but there was no association with cardiovascular (HR=1.00, 95% CI: 0.97-1.04) or respiratory mortality (HR=1.02, 95% CI: 0.96-1.07) (Table 3). Sex-specific analyses showed that the association with cancer-related mortality was prominent in the female subsample. As a *post-hoc* analysis, we further explored whether lung cancer (ICD-9: 162; ICD-10: C34) was the main driver of cancer-related mortality among females, which was confirmed (HR=1.11, 95% CI: 1.02-1.21). Among males, mortality due to natural cases other than cancer, cardiovascular or respiratory was significantly associated with early life exposure to PM_2.5_. Although not statistically significant, *post-hoc* we found increased risk of death (HR=1.08, 95% CI: 1.00-1.17; *p*=0.055) due to neurodegenerative disorders (Alzheimer’s disease and related disorders [ICD-9: 046.1, 290.0–290.4, 294, 331; ICD-10: F00-F03, G30-31], Parkinson’s disease [ICD-9: 332; ICD-10: G20-22], amyotrophic lateral sclerosis [ICD-9: 335.2; ICD-10: G12.2]).

**Table 3:** Associations between early life PM_2.5_ exposure and cause-specific mortality in Scottish Longitudinal Study Birth Cohort of 1936.

| Cause-specific mortality | Model 1<sup>a</sup><br>HR (95% CI) | Model 2<sup>b</sup> |  |  |
| --- | --- | --- | --- | --- |
|  |  | Total<br>HR (95% CI) | Female<br>HR (95% CI) | Male<br>HR (95% CI) |
| PM<sub>2.5</sub> in early life (10µg m<sup>-3</sup> increase) |  |  |  |  |
| All natural causes | **1.03 (1.01-1.05)** | **1.03 (1.01-1.05)** | 1.03 (1.00-1.06) | **1.04 (1.01-1.06)** |
| Cancer | **1.05 (1.02-1.08)** | **1.05 (1.02-1.08)** | **1.08 (1.03-1.13)** | 1.03 (0.99-1.07) |
| Cardiovascular | 1.00 (0.97-1.04) | 1.00 (0.97-1.04) | 0.98 (0.93-1.04) | 1.01 (0.97-1.06) |
| Respiratory | 1.02 (0.96-1.08) | 1.02 (0.96-1.07) | 1.02 (0.94-1.10) | 1.02 (0.94-1.11) |
| Other causes | **1.06 (1.01-1.10)** | **1.05 (1.01-1.10)** | 0.99 (0.91-1.08) | **1.08 (1.03-1.14)** |

Source: Scottish Longitudinal Study. The total sample size was $n=1947$ . Cox proportional hazard regressions were run with competing risks (i.e., outcome of interest versus other mortality) for each outcome separately. Hazard Ratios (HR) with 95% confidence intervals (CI) are presented. Bolded values are significant ( $p<0.05$ ). Cause-specific mortality was available between April 1991 and December 2017.
<sup>a</sup>Model 1 was adjusted for age and sex(strata).
<sup>b</sup>Model 2 was adjusted for age, (sex [strata],) parental occupational social position, mother's marital status, and mother's age.

The total effect of early life PM_2.5_ exposure on all-cause mortality was decomposed into direct association and indirect association mediated by childhood general cognitive ability. Individuals with ‘high’ early life PM_2.5_ exposure (≥50μg m^-3^) had a 39-month shorter median survival time in comparison to those from relatively ‘low’ concentration areas (<50μg m^-3^). Regression-based causal mediation analysis suggested that – in the fully adjusted model – being exposed to high concentrations of PM_2.5_ was associated with a 22% higher risk of all-cause mortality (95% CI: 1.08-1.38) (i.e., *total effects*). Approximately 25% of this association was mediated by general cognitive ability (HR=1.05; 95% CI: 1.03-1.07) (i.e., *indirect effects*): higher PM_2.5_ levels contributed to lower test scores, and lower test scores were associated with higher risk of all-cause mortality. The *direct effect* (i.e., not explained by general cognitive ability) of PM_2.5_ exposure on all-cause mortality was HR=1.17 (95% CI: 1.03-1.32) (Figure 4; Supplementary Table 2).

**Figure 4:**
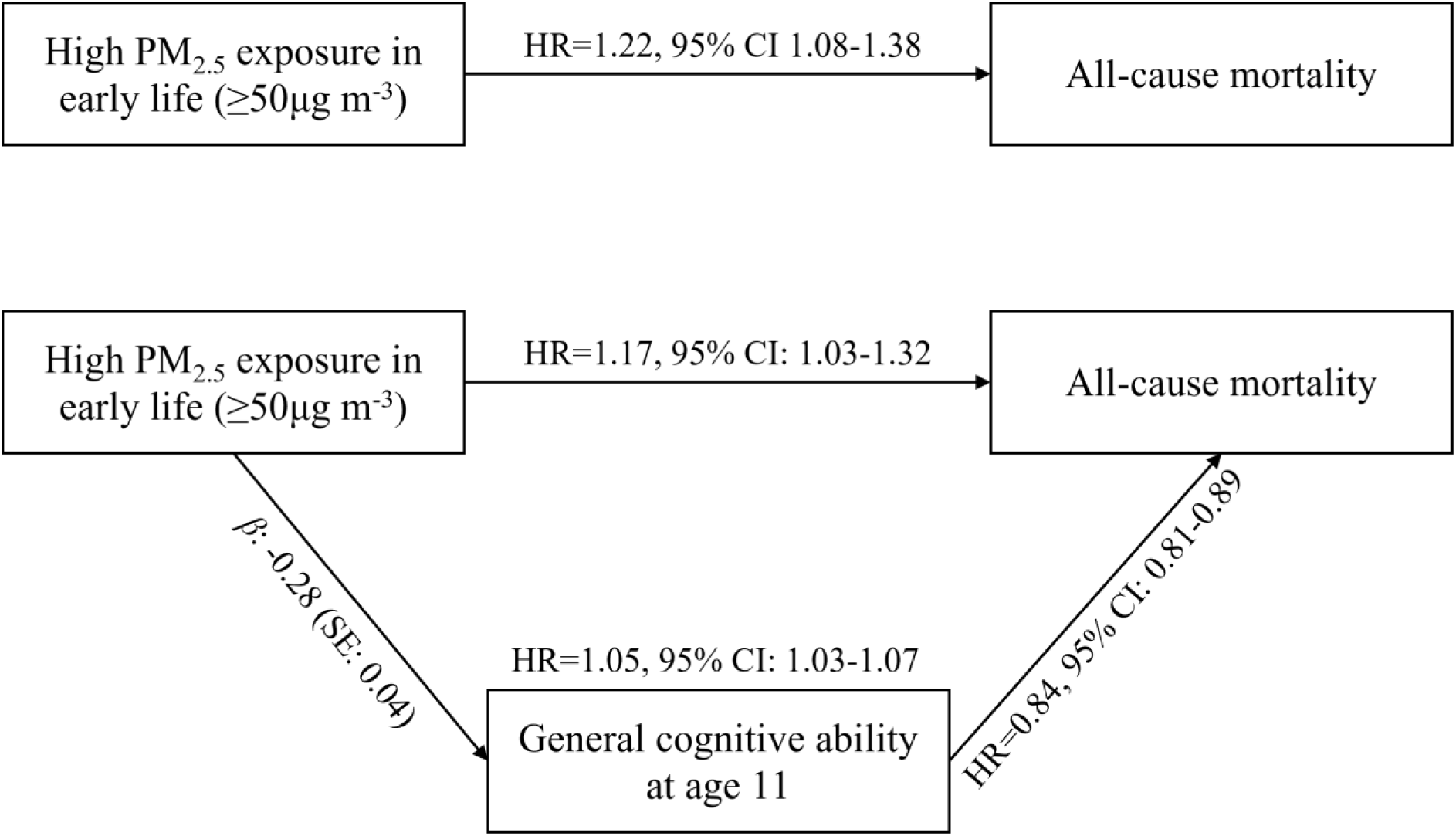
Total, direct, and indirect associations between ‘high’ early life PM_2.5_ exposure (≥50μg m^-3^) and all-cause mortality, with mediation through childhood general cognitive ability. All-cause mortality was available between June 1947 and June 2022. Model was adjusted for age, sex (strata), parental occupational social position, mother’s marital status, and mother’s age. The total sample size was *n*=2734. Source: Scottish Longitudinal Study.

Sensitivity analyses determined that 1-IQR increase in PM_2.5_ exposure was associated with 8% (95% CI: 1.03-1.13) increased risk of all-cause mortality in the fully adjusted model (Supplementary Table 3); findings for dichotomised air pollution levels (<50μg m^-3^, ≥50μg m^-3^) can be found in Supplementary Table 4. Adjusting for urban-rural differences in 1939 slightly attenuated the findings which did not remain significant in the female subsample (Supplementary Table 5). Imputing missing data through multiple imputation did not change the results (Supplementary Table 6); excluding participants with very high exposure (≥100μg m^-3^) only led to marginal changes (Supplementary Table 7). Exploring competing risks with Fine-Gray proportional subdistribution hazards regression produced the same cause-specific associations as presented earlier (Supplementary Table 8). Finally, after applying the 40μg m^-3^ cut-off for the mediation analysis, 21% of the total association was mediated by childhood cognitive ability (Supplementary Table 9; Supplementary Figure 1).

## DISCUSSION

Using a representative administrative cohort, we explored the direct and indirect associations between PM_2.5_ concentrations in early life and mortality across 75 years in 2734 participants. Exposure to higher levels of PM_2.5_ were associated with increased hazard of all-cause mortality, especially between the ages of 65 and 86. Competing risk analyses highlighted that associations were only present for cancer-related deaths, especially for lung-cancer mortality among females. One potential pathway connecting early life air pollution exposure to mortality is via lower childhood cognitive ability, which mediated 25% of the total associations in our sample.

This study strengthens the evidence base linking long-term PM_2.5_ exposure to increased risk of mortality in the UK and worldwide (2, 6–11). The estimated HR was 1.03 per 10μg m^-3^ incremental increase for all-cause mortality, which is much lower than the risk (Risk Ratio[RR]=1.08) estimated in a recent meta-analysis (2); however, this is not unexpected as previous findings with long-term follow-ups pointed towards decreasing effect sizes by more distal air pollution exposures in comparison to proximal ones (8, 46). Given the wide range of PM_2.5_ exposures, and the gentle, but steadily increasing slope of association, PM_2.5_ first crossed the HR=1 at the relatively higher concentration of 38μg m^-3^, which is much higher than thresholds of unhealthy exposure suggested by the literature (2, 47), including the WHO guidelines (48). We are not aware of previous investigations with a follow-up time of longer than 40 years (e.g., (8)) and making direct comparison to the literature is challenging; still, it is remarkable that toxic levels of air pollution might contribute to mortality 60-80 years after exposure.

Early life PM_2.5_ exposure was more strongly associated with all-cause mortality between age 65 and 86 and with cancer-related mortality, especially lung cancer. In contrary to all-cause mortality, HR for lung cancer in our female subsample (HR=1.11) was close to identical to risks reported in the literature based on shorter follow-up studies (RR=1.12) (2). Cancer-related mortality is the second leading cause of death according to the Global Burden of Disease Study, with tracheal, bronchus, and lung cancer being the leading cause of cancer deaths (49). Their relative proportion in comparison to other cancers is particularly high between the ages of 60 and 80 years (49), which overlaps with our findings. In contrary to the literature suggesting consistent associations between PM_2.5_ pollution and cardiovascular mortality (2), there was null association in our study. It might be plausible that PM_2.5_ in early life, or its chemical composition in the late 1930s, did not have such a long-lasting association with cardiovascular diseases or that early life is not a sensitive period for late adulthood cardiovascular events. Air pollution exposure has been also associated with neurodegenerative disorders in the literature (50), with stronger associations in males (51), which align with our preliminary findings. Still, these hypotheses cannot be answered from this study and need to be explored in further investigations.

During the prenatal period, air pollution-induced oxidative stress and systematic inflammation in the mother’s body may affect *in utero* development (52), and there is some evidence indicating that very tiny particles might even translocate into the placenta causing oxidative stress in the foetus (53). In early life, inhaled air pollutants can affect the children’s development more directly (17, 54). Toxic environmental exposures – through oxidative stress and inflammation – are associated with molecular changes in the somatic cells, and these alterations might survive subsequent cell replications and affect healthy development and longevity. For example, modified DNA methylation patterns, responsible for gene expression and genome stability, as well as shorter telomeres, a marker of cellular ageing, have been associated with air pollution exposure in early and later life (17, 55). A recent Scottish study, based on a sample (Lothian Birth Cohort 1936) selected from the same 1936-born population as in our study, showed that higher air pollution exposure around birth was associated with shorter DNA methylation-based telomere length in late adulthood, especially among females (12), even after adjusting for a variety of life-course social and behavioural (e.g. smoking) risk factors. Epigenetic markers increase the risk of developing cancer (56) and mortality (57), presenting a plausible explanation for our (sex-specific) findings.

Air pollution in early life may also affect brain development: maternal air pollution exposure from the 25^th^ gestational week onwards is associated with head growth (58) and being exposed to higher prenatal and postnatal PM_2.5_ concentration is linked to poorer cognitive performance in childhood (52, 59, 60). The human brain undergoes rapid developments in the early years which continues well into childhood with the differentiation and maturation of neurons and myelination of axons (61, 62). A whole population follow up on the SMS1947 with more than 65000 participants showed that higher general cognitive ability scores in childhood decreased all-cause mortality and cause-specific mortality for almost all major causes of death, including smoking-related cancers (18). Higher cognitive ability in childhood is a powerful predictor of longevity with pathways including higher educational attainment and adult socioeconomic status, which can impact among others health literacy, health behaviour, and diseases management (19, 63). Our findings adds that higher early life PM_2.5_ exposure modestly decreases general cognitive ability in childhood (even after adjusting for parental occupational social position), thus indirectly contributing to all-cause mortality; approximately one-quarter of the total association was mediated through childhood cognitive ability presenting an important pathway between early life toxic exposure and mortality.

### Strengths and limitations

This study demonstrated the feasibility of exploring the relationship between early life PM_2.5_ exposure and mortality over almost the entire life course. The study benefitted from high quality data linkages between different historical and contemporary administrative sources, included key family and household confounders in early childhood, a validated measure of general cognitive ability at age 11, and an exceptionally long follow-up of mortality stretching over 75 years. Residential information at age 3 was derived from administrative data, thus not prone to recall bias. However, several key limitations should be considered when interpreting the results. First, cause-specific mortality was only available for a relatively shorter follow-up time (∼26 years). Although we do not assume that longer follow-up would have change the reported associations with cancer-related deaths, it is plausible that adding further mortality records across the life course may reveal other cause-specific mortality associations in future studies. Second, information on mortality before age 11 was not available in the SLSBC1936. Given the high mortality rates among 1-year olds in England during the 1930s (64), and the strong association between PM_2.5_ concentrations and infant mortality in high pollution contexts (e.g. India (65)), it is plausible that air pollution had a strong effect on mortality in the earlier years which we were not able to capture. Third, early life residential location was only available in the 1939 National Identity Register. Although the probability of residential mobility for this birth cohort is relatively low in their earliest years (66), we cannot ascertain whether air pollution exposure during prenatal or during postnatal periods contributed to higher risk of mortality. Moreover, missing residential history during childhood introduces further uncertainty to identify sensitive (or critical) exposure windows. Fourth, there are inherent uncertainties at all stages of the emissions estimation process, even for the present day, and many of these problems are magnified when trying to recreate a historical context. The same atmospheric chemistry transport models for newer modelling years (2009-2010) have been evaluated against concurrent PM_2.5_ concentrations, which indicated good agreement between estimated and measured concentrations, but also suggested that PM_2.5_ concentrations might be underestimated in the EMEP4UK models (30). While there are increasing uncertainties when pollutants are estimated further back in time, their relative spatial concentrations can be considered as sufficiently accurate. Last, key air pollution regulatory policies throughout the 20^th^ century (e.g., Clean Air Acts 1956, 1968, 1993) reduced PM_2.5_ concentrations in the UK (67), and probably changed its chemical components. Pollutant concentrations experienced by sample participants in their early lives are rarely present in high-income countries anymore. Still, with increasing global inequities in pollution though raising concentration in low– and middle-income countries (48), our findings have relevant global implications.

Future studies could capitalise on routinely collected data utilising census responses, healthcare service use data and Vital Events on marriages and children’s birth to extract information on residential locations across the entire life course and provide a more comprehensive assessment on how air pollution contributes to morbidity and mortality from foetal period onwards. Mechanistic pathways, including social, cognitive, and biological processes, should be investigated which have the potential to connect childhood exposure to late adulthood health.

## Conclusions

Exposure to higher PM_2.5_ in early life was associated with higher all-cause and cancer-related mortality, especially between the ages of 65 and 75, with one-quarter of the association being mediated through lower childhood general cognitive ability. Future investigation should explore mechanistic pathways and assess the relative importance of air pollution exposure at different time-points in our lives. The findings suggest that air pollution may have very long-term impact on health and longevity, with its detrimental impact lasting long after mitigation policies reduced concentration levels.

## Data Availability

Due to the sensitive nature of the data produced for this study, it is only accessible for SLS Approved Researchers from a Safe Setting. Access to data is granted after the SLS Research Board's approval.

## ACKNOWLEDGEMENT

The help provided by staff of the Longitudinal Studies Centre – Scotland (LSCS) is acknowledged. The LSCS is supported by the ESRC/JISC, the Scottish Funding Council, the Chief Scientist’s Office and the Scottish Government. The authors alone are responsible for the interpretation of the data. Census output is Crown copyright and is reproduced with the permission of the Controller of HMSO and the King’s Printer for Scotland.

## Supplementary Materials

**Supplementary Table 1:** Model fit with penalised splines

| Degrees of freedom | Bayesian Information Criterion |  |  |
| --- | --- | --- | --- |
|  | Total | Female | Male |
| Two | 21040.84 | 9350.212 | 11722.6 |
| Three | 21047.21 | 9356.144 | 11727.24 |
| Four | 21053.36 | 9362.217 | 11732.27 |

Source: Scottish Longitudinal Study. The sample size was $n=2734$ . Cox proportional hazard regression were fitted with penalised splines with differing degrees of freedom to explore concentration-response relationship. Models were adjusted for age, (sex (strata),) parental occupational social position, mother's marital status, and mother's age. All-cause mortality was available between June 1947 and June 2022.

**Supplementary Table 2:** Outcome and mediator models in the regression-based causal mediation analysis (50μg m^-3^ cut-off)

| Covariates | Mediator model |  | Outcome model |  |
| --- | --- | --- | --- | --- |
| | $\beta$ (SE) | p-value | HR (95% CI) | p-value |
| High PM<sub>2.5</sub> in early life ($\geq 50\mu\text{g m}^{-3}$) | -0.28 (0.05) | <0.001 | 1.17 (1.04-1.31) | 0.011 |
| Age (per 1 SD increase) | 0.13 (0.02) | <0.001 | 1.03 (0.98-1.08) | 0.196 |
| Sex |  |  |  |  |
| Male | ref |  | ref |  |
| Female | 0.13 (0.04) | <0.001 | 0.66 (0.60-0.73) | <0.001 |
| Parental occupational social position (per 1 SD increase) | 0.25 (0.02) | <0.001 | 0.95 (0.90-1.01) | 0.081 |
| Mother's marital status |  |  |  |  |
| Married | ref |  | ref |  |
| Not married | -0.22 (0.10) | 0.029 | 1.18 (0.90-1.55) | 0.237 |
| Mother's age in 1939 (per 1 SD increase) | -0.02 (0.02) | 0.297 | 0.96 (0.91-1.00) | 0.070 |
| Moray House test score (per 1 SD increase) | NA |  | 0.85 (0.81-0.89) | <0.001 |

Source: Scottish Longitudinal Study. The sample size was $n=2734$ . Exposure was dichotomised between low ( $<50\mu\text{g m}^{-3}$ ) and high ( $\geq 50\mu\text{g m}^{-3}$ ), separating approximately 80% and 20% of the sample. Cox proportional hazard regression was fitted for the outcome model (i.e., Hazard Ratios [HR] with 95% confidence intervals [CI]), linear regression was fitted for the mediator model (i.e., standardized coefficients with standard errors [SE]). All-cause mortality was available between June 1947 and June 2022. Abbreviations: NA=not applicable; SD=standard deviation.

**Supplementary Table 3:** Associations between early life PM_2.5_ exposure and all-cause mortality expressed as interquartile range change.

| Exposure | Model 1<sup>a</sup><br>HR (95% CI) | Model 2<sup>b</sup> |  |  |
| --- | --- | --- | --- | --- |
|  |  | Total<br>HR (95% CI) | Female<br>HR (95% CI) | Male<br>HR (95% CI) |
| *a) During total follow-up* |  |  |  |  |
| PM<sub>2.5</sub> in early life (IQR increase) | **1.08 (1.04-1.13)** | **1.08 (1.03-1.13)** | 1.09 (1.02-1.17) | **1.07 (1.01-1.13)** |
| *b) By follow-up intervals<sup>a</sup>* |  |  |  |  |
| PM<sub>2.5</sub> in early life (IQR increase) |  |  |  |  |
| Age 11-55 year | 1.01 (0.90- 1.14) | 1.01 (0.90-1.13) | 1.02 (0.83-1.24) | 1.00 (0.87-1.16) |
| Age 55-65 year | 1.03 (0.92-1.15) | 1.02 (0.91-1.14) | 1.15 (0.96-1.39) | 0.96 (0.83-1.11) |
| Age 65-75 year | **1.15 (1.06-1.24)** | **1.14 (1.05-1.23)** | 1.05 (0.92-1.20) | **1.20 (1.09-1.33)** |
| Age 75-86 year | **1.08 (1.01-1.16)** | **1.08 (1.01-1.16)** | **1.11 (1.02-1.22)** | 1.05 (0.96-1.16) |

Source: Scottish Longitudinal Study. The sample size was $n=2734$ . Cox proportional hazard regression were fitted and Hazard Ratios (HR) with 95% confidence intervals (CI) are presented. Bolded values are significant ( $p<0.05$ ). All-cause mortality was available between June 1947 and June 2022. Abbreviation; IQR=interquartile range.
<sup>a</sup> Model 1 was adjusted for age and sex.
<sup>b</sup> Model 2 was adjusted for age, (sex (strata),) parental occupational social position, mother's marital status, and mother's age.

**Supplementary Table 4:** Associations between low versus high early life PM_2.5_ exposure and all-cause mortality.

| Exposure | Model 1<sup>a</sup><br>HR (95% CI) | Model 2<sup>b</sup> |  |  |
| --- | --- | --- | --- | --- |
|  |  | Total<br>HR (95% CI) | Female<br>HR (95% CI) | Male<br>HR (95% CI) |
| *a) During total follow-up* |  |  |  |  |
| High PM<sub>2.5</sub> in early life (≥50μg m<sup>-3</sup>) | **1.23 (1.09- 1.38)** | **1.22 (1.09-1.37)** | **1.21 (1.01- 1.45)** | **1.23 (1.05- 1.44)** |
| *b) By follow-up intervals<sup>a</sup>* |  |  |  |  |
| High PM<sub>2.5</sub> in early life (≥50μg m<sup>-3</sup>) |  |  |  |  |
| Age 11-55 year | 1.02 (0.74-1.41) | 1.01 (0.73-1.39) | 0.87 (0.49-1.54) | 1.09 (0.73-1.61) |
| Age 55-65 year | 1.06 (0.78-1.45) | 1.05 (0.77-1.43) | 1.26 (0.74-2.13) | 0.96 (0.65-1.41) |
| Age 65-75 year | **1.47 (1.18-1.82)** | **1.46 (1.17-1.81)** | 1.17 (0.82-1.66) | **1.70 (1.29-2.24)** |
| Age 75-86 year | **1.22 (1.02-1.46)** | **1.22 (1.02-1.46)** | **1.31 (1.02-1.69)** | 1.13 (0.87-1.48) |

Source: Scottish Longitudinal Study. The sample size was $n=2734$ . Exposure was dichotomised between low ( $<50\mu\text{g m}^{-3}$ ) and high ( $\geq 50\mu\text{g m}^{-3}$ ), separating approximately 80% and 20% of the sample. Cox proportional hazard regression were fitted and Hazard Ratios (HR) with 95% confidence intervals (CI) are presented. Bolded values are significant ( $p<0.05$ ). All-cause mortality was available between June 1947 and June 2022.
<sup>a</sup> Model 1 was adjusted for age and sex.
<sup>b</sup> Model 2 was adjusted for age, (sex (strata),) parental occupational social position, mother's marital status, and mother's age.

**Supplementary Table 5:** Associations between early life PM_2.5_ exposure and all-cause mortality a) during the total follow up and b) by follow-up intervals after adjusting for residential location.

| Exposure | Model 1<sup>a</sup><br>HR (95% CI) | Model 2<sup>b</sup> |  |  |
| --- | --- | --- | --- | --- |
|  |  | Total<br>HR (95% CI) | Female<br>HR (95% CI) | Male<br>HR (95% CI) |
| *a) During total follow-up* |  |  |  |  |
| PM<sub>2.5</sub> in early life (10µg m<sup>-3</sup> increase) | **1.03 (1.01-1.05)** | **1.03 (1.01-1.05)** | 1.02 (0.99-1.05) | **1.04 (1.01-1.07)** |
| *b) By follow-up intervals<sup>a</sup>* |  |  |  |  |
| PM<sub>2.5</sub> in early life (10µg m<sup>-3</sup> increase) |  |  |  |  |
| Age 11-55 year | 1.01 (0.97- 1.05) | 1.00 (0.96- 1.05) | 0.99 (0.92-1.06) | 1.02 (0.96-1.07) |
| Age 55-65 year | 1.01 (0.97- 1.05) | 1.01 (0.97- 1.05) | 1.03 (0.97-1.10) | 1.00 (0.95-1.05) |
| Age 65-75 year | **1.05 (1.02- 1.08)** | **1.05 (1.02- 1.08)** | 1.00 (0.96-1.05) | **1.08 (1.04-1.12)** |
| Age 75-86 year | **1.03 (1.00- 1.06)** | **1.03 (1.00- 1.06)** | 1.02 (0.99-1.06) | 1.03 (0.99-1.07) |

Source: Scottish Longitudinal Study. The sample size was $n=2734$ . Cox proportional hazard regression were fitted and Hazard Ratios (HR) with 95% confidence intervals (CI) are presented. Bolded values are significant ( $p<0.05$ ). All-cause mortality was available between June 1947 and June 2022.
<sup>a</sup> Model 1 was adjusted for age, sex and residential location (urban, rural).
<sup>b</sup> Model 2 was adjusted for age, (sex (strata),) residential location (urban, rural), parental occupational position, mother's marital status, and mother's age.

**Supplementary Table 6:** Associations between early life PM_2.5_ exposure and all-cause mortality a) during the total follow up and b) by follow-up intervals after multiple imputation.

| Exposure | Model 1<sup>a</sup><br>HR (95% CI) | Model 2<sup>b</sup> |  |  |
| --- | --- | --- | --- | --- |
|  |  | Total<br>HR (95% CI) | Female<br>HR (95% CI) | Male<br>HR (95% CI) |
| *a) During total follow-up* |  |  |  |  |
| PM<sub>2.5</sub> in early life (10µg m<sup>-3</sup> increase) | **1.03 (1.02-1.04)** | **1.03 (1.01-1.04)** | **1.02 (1.00-1.04)** | **1.03 (1.01-1.05)** |
| *b) By follow-up intervals<sup>a</sup>* |  |  |  |  |
| PM<sub>2.5</sub> in early life (10µg m<sup>-3</sup> increase) |  |  |  |  |
| Age 11-55 year | 1.01 (0.98-1.04) | 1.01 (0.98-1.05) | 1.02 (0.97-1.08) | 1.00 (0.96-1.05) |
| Age 55-65 year | 1.01 (0.98-1.05) | 1.01 (0.97-1.04) | 1.04 (0.98-1.09) | 0.99 (0.95-1.03) |
| Age 65-75 year | **1.04 (1.01-1.06)** | **1.04 (1.01-1.06)** | 1.02 (0.98-1.06) | **1.05 (1.02-1.08)** |
| Age 75-86 year | **1.03 (1.01-1.05)** | **1.03 (1.01-1.05)** | **1.04 (1.01-1.07)** | 1.02 (0.99-1.05) |

Source: Scottish Longitudinal Study. The sample size was $n=3626$ . After imputing missing data with multiple imputation by chained equation (24-25 imputed dataset depending on the model; imputations were based on all covariates including childhood general cognitive ability), Cox proportional hazard regression were fitted and Hazard Ratios (HR) with 95% confidence intervals (CI) are presented. Bolded values are significant ( $p<0.05$ ). All-cause mortality was available between June 1947 and June 2022.
<sup>a</sup> Model 1 was adjusted for age and sex.
<sup>b</sup> Model 2 was adjusted for age, (sex (strata),) parental occupational position, mother's marital status, and mother's age.

**Supplementary Table 7:** Associations between early life PM_2.5_ exposure and all-cause mortality a) during the total follow up and b) by follow-up intervals after dropping individuals with very high exposure (≥100μg m^-3^).

| Exposure | Model 1<sup>a</sup><br>HR (95% CI) | Model 2<sup>b</sup> |  |  |
| --- | --- | --- | --- | --- |
|  |  | Total<br>HR (95% CI) | Female<br>HR (95% CI) | Male<br>HR (95% CI) |
| *a) During total follow-up* |  |  |  |  |
| PM<sub>2.5</sub> in early life (10μg m<sup>-3</sup> increase) | **1.03 (1.01-1.05)** | **1.03 (1.01-1.05)** | **1.03 (1.00-1.07)** | 1.02 (0.99-1.06) |
| *b) By follow-up intervals<sup>a</sup>* |  |  |  |  |
| PM<sub>2.5</sub> in early life (10μg m<sup>-3</sup> increase) |  |  |  |  |
| Age 11-55 year | 1.02 (0.96-1.08) | 1.02 (0.96-1.08) | 1.01 (0.91-1.11) | 1.03 (0.95-1.11) |
| Age 55-65 year | 1.01 (0.95-1.07) | 1.00 (0.95-1.07) | 0.98 (0.88-1.09) | 1.02 (0.95-1.09) |
| Age 65-75 year | **1.06 (1.01-1.10)** | **1.05 (1.01-1.10)** | 1.03 (0.97-1.10) | **1.07 (1.02-1.14)** |
| Age 75-86 year | 1.02 (0.99-1.06) | 1.02 (0.99-1.06) | **1.05 (1.01-1.10)** | 0.98 (0.93-1.04) |

Source: Scottish Longitudinal Study. The sample size was $n=2487$ . After imputing missing data with multiple imputation by chained equation (24-25 imputed dataset depending on the model), Cox proportional hazard regression were fitted and Hazard Ratios (HR) with 95% confidence intervals (CI) are presented. Bolded values are significant ( $p<0.05$ ). All-cause mortality was available between June 1947 and June 2022.
<sup>a</sup> Model 1 was adjusted for age and sex.
<sup>b</sup> Model 2 was adjusted for age, (sex (strata),) parental occupational position, mother's marital status, and mother's age.

**Supplementary Table 8:**
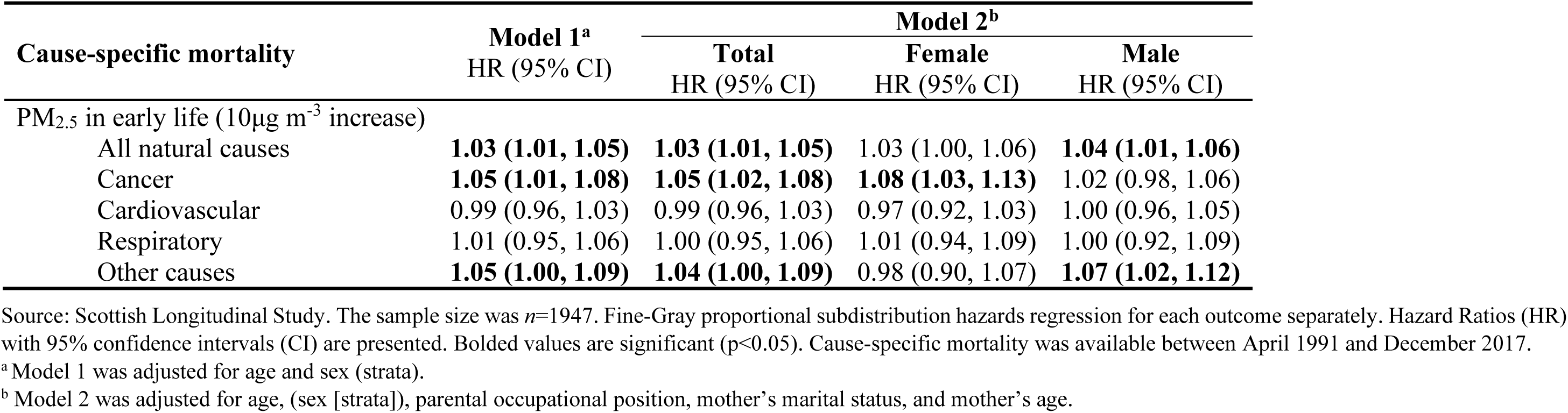
Associations between early life PM_2.5_ exposure and cause-specific mortality calculated with Fine-Gray regression.

| Cause-specific mortality | Model 1<sup>a</sup><br>HR (95% CI) | Model 2<sup>b</sup> |  |  |
| --- | --- | --- | --- | --- |
|  |  | Total<br>HR (95% CI) | Female<br>HR (95% CI) | Male<br>HR (95% CI) |
| PM<sub>2.5</sub> in early life ($10\mu\text{g m}^{-3}$ increase) | | | | |
| All natural causes | **1.03 (1.01, 1.05)** | **1.03 (1.01, 1.05)** | 1.03 (1.00, 1.06) | **1.04 (1.01, 1.06)** |
| Cancer | **1.05 (1.01, 1.08)** | **1.05 (1.02, 1.08)** | **1.08 (1.03, 1.13)** | 1.02 (0.98, 1.06) |
| Cardiovascular | 0.99 (0.96, 1.03) | 0.99 (0.96, 1.03) | 0.97 (0.92, 1.03) | 1.00 (0.96, 1.05) |
| Respiratory | 1.01 (0.95, 1.06) | 1.00 (0.95, 1.06) | 1.01 (0.94, 1.09) | 1.00 (0.92, 1.09) |
| Other causes | **1.05 (1.00, 1.09)** | **1.04 (1.00, 1.09)** | 0.98 (0.90, 1.07) | **1.07 (1.02, 1.12)** |

Source: Scottish Longitudinal Study. The sample size was $n=1947$ . Fine-Gray proportional subdistribution hazards regression for each outcome separately. Hazard Ratios (HR) with 95% confidence intervals (CI) are presented. Bolded values are significant ( $p<0.05$ ). Cause-specific mortality was available between April 1991 and December 2017.
<sup>a</sup> Model 1 was adjusted for age and sex (strata).
<sup>b</sup> Model 2 was adjusted for age, (sex [strata]), parental occupational position, mother's marital status, and mother's age.

**Supplementary Table 9:** Outcome and mediator models in the regression-based causal mediation analysis (40μg m^-3^ cut-off)

| Covariates | Mediator model |  | Outcome model |  |
| --- | --- | --- | --- | --- |
| | $\beta$ (SE) | p-value | HR (95% CI) | p-value |
| High PM<sub>2.5</sub> in early life ($\geq 40\mu\text{g m}^{-3}$) | -0.17 (0.04) | <0.001 | 1.13 (1.01-1.26) | 0.038 |
| Age (per 1 SD increase) | 0.12 (0.02) | <0.001 | 1.03 (0.99-1.09) | 0.175 |
| Sex |  |  |  |  |
| Male | ref |  | ref |  |
| Female | 0.13 (0.04) | <0.001 | 0.66 (0.60-0.73) | <0.001 |
| Parental occupational social position (per 1 SD increase) | 0.25 (0.02) | <0.001 | 0.95 (0.90-1.01) | 0.085 |
| Mother's marital status |  |  |  |  |
| Married | ref |  | ref |  |
| Not married | -0.22 (0.10) | 0.031 | 1.18 (0.90-1.55) | 0.233 |
| Mother's age in 1939 (per 1 SD increase) | -0.02 (0.02) | 0.268 | 0.96 (0.91-1.01) | 0.083 |
| Moray House test score (per 1 SD increase) | NA |  | 0.85 (0.80-0.89) | <0.001 |

Source: Scottish Longitudinal Study. The sample size was $n=2734$ . Exposure was dichotomised between low ( $<40\mu\text{g m}^{-3}$ ) and high ( $\geq 40\mu\text{g m}^{-3}$ ), separating approximately 75% and 25% of the sample. Cox proportional hazard regression was fitted for the outcome model (i.e., Hazard Ratios [HR] with 95% confidence intervals [CI]), linear regression was fitted for the mediator model (i.e., standardized coefficients [ $\beta$ ] with standard errors [SE]). All-cause mortality was available between June 1947 and June 2022. Abbreviations: NA=not applicable; SD=standard deviation.

**Supplementary Figure 1:**
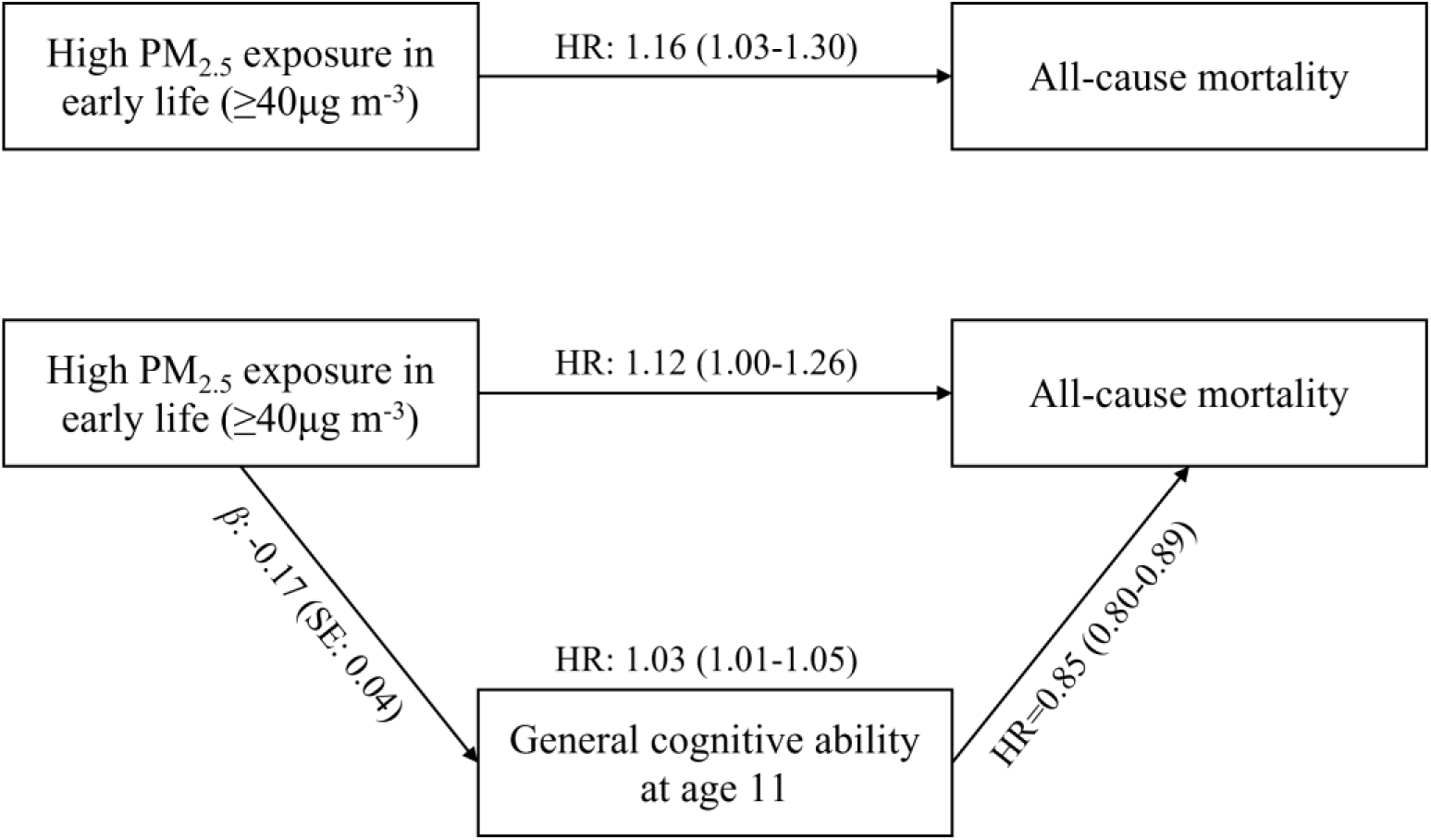
Total, direct, and indirect associations between high PM_2.5_ exposure in early life (≥40μg m-3) and all-cause mortality, with mediation through childhood general cognitive ability. All-cause mortality was available between June 1947 and June 2022. Model was adjusted for age, sex (strata), parental occupational position, mother’s marital status, and mother’s age. The sample size was *n*=2734. Source: Scottish Longitudinal Study.

